# Investigation Profiles and Costs of Preoperative Evaluation among Vascular Surgery Patients at a Tertiary Hospital in Addis Ababa, Ethiopia

**DOI:** 10.64898/2025.12.28.25343111

**Authors:** Karenzi Irenee David, Tamiru Deneke, Abigael Abiy Mesfin, Samuel Zerihun Tesfaye, Dawit Gebregiorgis

## Abstract

**Background:** Preoperative evaluation is essential for ensuring patient safety prior to vascular surgery. However, overutilization of routine tests without clinical indication can impose significant financial burden on people, especially those who are living in resource-limited settings. This study aim was to assess preoperative evaluation in vascular patients with its associated direct patient cost and cost drivers in Tikur Anbessa Specialized Hospital (TASH).

**Objective:** To assess the direct cost, and key cost drivers of preoperative investigations among patients who underwent elective vascular surgery at Tikur Anbessa Specialized Hospital (TASH) from Jan2022 to December 2024.

**Methods:** An institutional-based retrospective cross-sectional study was conducted for vascular patients operated from January 2022 to December 2024 at TASH , assessing costs associated with preoperative investigations. Data were collected from medical records, and finance logs. Investigations were categorized as indicated or unnecessary based on National Institute for Health and Care Excellence (NICE) guidelines 2016. Costs were determined using TASH service fees and private center rates and patients’ information. Data were analyzed using SPSS version 21.

**Results:** A total of 165 elective vascular surgery patients were randomly selected; the mean age of participants was 49.6 years, with 53.3% male and 46.7% female. Peripheral artery disease was the most common diagnosis (47.3%). The average hospital stay was 13.7 days. The mean total cost of preoperative investigations per patient was 9,717.40 Ethiopian Birrs (ETB), with imaging averaging 8,797 ETB and laboratory tests averaging 1,328 ETB. HIV screening was not commonly provided for elective vascular patients. Linear Regression was conducted to explore relationships between patient demographics, clinical factors, and both the number and cost of investigations. Age, ASA class, comorbidity and longer hospital days were positively associated with increased total cost.

**Conclusion:** Preoperative evaluation in vascular surgery at TASH is characterized by excessive use of routine and advanced investigations, some of which lack clinical indication. These practices contribute to substantial patient expenses, primarily through out of pocket payments. There is also gap in providing some investigation tests like serum albumin and calcium. Developing and implementing evidence based testing guidelines could limit the cost.

## Introduction

Preoperative evaluation or preoperative check-up is the prerequisite for preparing a patient for surgery. It is described as the clinical evaluation step that precedes the administration of anesthetics for surgery and other procedures. Data from many sources, including preoperative tests, must be considered. (1) Preoperative evaluation may include routine tests and indicated tests. The American Society of Anesthesiologists (ASA) Task Force on Preanesthesia Checkup (PAC) defines routine tests as those performed in the absence of any specific clinical indication or purpose, whereas indicated tests are those performed for specific clinical purposes or indications, such as confirming a clinical diagnosis or assessing the severity and progression of disease.

Patients with peripheral arterial disease have an increased risk of major cardiovascular complications such as myocardial infarction, heart failure, arrhythmias and stroke. For this reason, the preoperative evaluation aims to reduce mortality related to those complications, even though there is no standardized approach for the preoperative workup of those patients.(2) Preoperative cardiac evaluation and risk stratification in surgical patients, especially in vascular surgery, should be performed by any practicing surgeon. Owing to the systemic nature of atherosclerosis and similarities in pathophysiology and risk factors for both vascular and cardiac disease processes, most vascular patients suffer from cardiac disease(3,4). Preoperative laboratory investigations are considered important elements of preanaesthesia evaluation to determine the fitness for anesthesia and surgery. A few decades ago, this practice was closely surveyed because of its low input and high added cost. It was found that performing routine screening tests in otherwise healthy patients is of little value in detecting diseases and in changing anesthetic management or outcomes. Many investigations are costly, often detect minor abnormalities of no clinical relevance, may be risky to patients, and may cause unnecessary delay or cancellation of surgery. Selective testing reduces cost without sacrificing the safety or quality of surgical care.(5)

Advances in technology and the introduction of multiple laboratory tests a few decades ago led to an increase in the number of ordered tests, suggesting that an asymptomatic diagnosis will optimize care and reduce medical costs, but more testing can lead to an increased number of abnormal results, leading to additional unnecessary workups with increased medical costs and the possibility of increasing morbidity from more medical interventions. Preoperative testing should be based on a targeted history and physical examination for patient comorbidities and types of surgery planned.(6)

Vascular surgery procedures are grouped into three categories according to the likelihood of developing perioperative major adverse cardiac events (MACEs): high-risk procedures with risk >5%, intermediate-risk procedures (1%-5%) and low-risk procedures with risk <1%. Preoperative cardiac evaluation is based on the abovementioned categories.(7) Effective preoperative evaluation of vascular patients includes cardiac, respiratory system, renal, metabolic, coagulation, and blood group data and cross matching and specialist input. The prevalence of cardiovascular disease in vascular patients is 25–60%, that of smoking is 35–85%, and the prevalence of respiratory disease is 10–50% in vascular patients. The prevalence of renal system dysfunction is 4–13%. (8) There are large variations in the length of hospital stay, mainly due to practice patterns rather than the severity of disease, and interventions that can decrease the length of hospital stay can result in cost savings.(9) Economic analysis suggests that preoperative workup guidelines can reduce the cost of preoperative investigations(10).

Many patients with cardiovascular diseases need surgical treatment even if they present with many comorbidities that may predispose them to perioperative adverse events. Preoperative preparation of these patients requires different clinical investigations that might increase the cost of care, and for some patients, safe and timely surgery may not be affordable. To the best of our knowledge, the total cost of preoperative investigations and their key cost drivers for vascular patients receiving surgical care at Tikur Anbessa Specializzed Hospital (TASH) have not been studied. Understanding the cost related to preoperative investigations for vascular patients and unnecessary increments will lead to the implementation of evidence-based guidelines with the aim of enhancing the cost-effectiveness of vascular surgery care at TASH.

This study assessed the total direct cost incurred by patients for preoperative investigations prior to undergoing vascular surgery. Specifically, this study sought to characterize the profile of preoperative investigations commonly performed for these patients and estimate the total and average direct costs related to the abovementioned investigations at Tikur Anbessa Specialized Hospital (TASH) and private health facilities. Additionally, this study provides insight into the routine preoperative tests performed and their associated financial burden on patients and the key cost drivers of preoperative investigations of vascular patients.

## Methods and Materials

### Study Design

This was an institutionally based retrospective cross-sectional study that assessed the profile of preoperative investigations in vascular patients and cost and key cost drivers related to investigations of vascular patients in the vascular surgery unit of TASH.

### Study setting

The study was conducted in TASH, which is the Ethiopian largest specialized and referral public hospital and is the main referral center for vascular surgery in the country. In 1998, the hospital was given to Addis Ababa University by the Ministry of Health to serve as the main teaching hospital. The hospital offers diagnosis and treatment for >500000 patients per year (32). It receives patients referred from across the country. TASH was selected for this study because it is the largest tertiary referral hospital and main referral center for vascular surgery in Ethiopia. This hospital is selected because it is the largest tertiary referral hospital and main referral center for vascular surgery in Ethiopia. Special tests will be requested before surgery. The above tests are performed in the hospital, but a portion of them are performed in private centers if they are not available in TASH.

In TASH, patients pay either through health insurance or out-of-pocket money, but in private centers, all pay out-of-pocket money.

### Source population

All vascular patients were admitted with confirmed surgical diagnoses and underwent preoperative evaluations in their medical records.

### Study population

Patients who were admitted with a confirmed surgical diagnosis and underwent preoperative evaluation from Jan 2022 to December 2024

Patients referred for vascular surgery are evaluated, investigated and planned for surgery in a referral clinic by a surgical team led by a consultant vascular surgeon. Some of the patients admitted for elective surgery are evaluated by an anesthesia team in the preanesthesia clinic on the day of admission. All patients were also evaluated by an anesthesia team in the surgical ward on the day before the operation. A comprehensive history and physical examination were performed for every patient, and imaging, laboratory tests and other special tests were requested and recorded in patient files before surgery. The above tests were performed in the hospital, but a portion of them were performed in private centers if they were not available in TASH. In hospitals, patients pay through health insurance coverage or out-of-pocket money; however, in private centers, all pay out-of-pocket money.

### Inclusion and Exclusion Criteria

#### Inclusion criteria

All patients were admitted and evaluated for elective surgery in the vascular unit from 2022--2024. All information regarding preoperative evaluation and the direct medical costs that patients paid in TASH and private health facilities for preoperative investigations was collected.

#### Exclusion criteria

- Patients in the abovementioned population whose data from preoperative investigations were missing in files and electronic medical records were excluded from the study.
- Patients who underwent surgery on an emergency basis without time for preoperative investigations were excluded from the study.
- Patients who underwent surgery as day cases.

#### Sample size determination

Data from the registry revealed 240 cases as the total population of patients operated on in the vascular unit per year. The acceptable precision for this research is 0.05. With these values, the sample size was calculated via the Yamane formula: 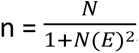, where n is the sample size, N is the finite population, and E is the level of significance (0.05 for this study). Accordingly, the sample size was 165, including a 10% nonresponse rate.

#### Sampling Technique

A random sampling technique in Microsoft Excel was used to recruit patients for this study. The retrieved patient medical record numbers (MRNs) were entered into an Excel sheet, a random list was generated, and 165 patients’ MRNs were obtained randomly. For each MRN, the corresponding file was retrieved from the hospital archive; if the file was not found, the next MRN on the random list was taken to replace the missing data.

#### Data collection and quality assurance

The data collection tool was developed based on similar studies and it was tested. Patients’ details and clinical data were retrieved from operating theater logbook on January 30th 2025 for research purposes. Patient details and clinical data were retrieved from the operating theater logbook, and patient medical records and cost information were collected from hospital service fee records. For tests performed in private centers, providing centers were contacted to obtain information on the cost of each investigation. The data were kept in password-protected Excel form and analyzed via SPSS version 21. This study used fully de-identified data collected from existing records. Authors did not access identifiable participant information during or after data collection.

Evidence-based recommendations for ordering preoperative investigations for elective surgery were used for each investigation and to denote indicated and non-indicated tests for a given procedure. The 2016 National Institute of Care for Excellence (NICE) guidelines (United Kingdom) was referred to as the recently updated guidelines for the preoperative investigation of non-cardiac surgery.

For every recruited patient, the direct cost (patient cost) of the investigation and the duration of hospital stay related to planned surgery were calculated. The data collection team was recruited and trained on the research protocol to maximize data quality. The data collection team comprised health professionals only. A predesigned data collection form was used to collect all needed information, and the data were entered into an Excel dataset for data cleaning before analysis.

### Study variables

#### Dependent Variables

Total direct cost of preoperative investigation

#### Independent Variable

- Age
- Sex
- ASA classification
- Admission Diagnosis
- Type of operation:
- Having comorbidities (diabetes mellitus, hypertension, heart disease, pulmonary disease, other)
- Smoking status: previous smoker, active smoker
- Alcohol use
- Investigation tests

#### Ethical Considerations

Ethical approval for this study was obtained from the Addis Ababa University, College of Health Sciences Institutional Review Board under Ref. No. DOS/REC/143/2025/2027. The study was conducted in accordance with the principles of the Declaration of Helsinki. Informed electronic consent was obtained from all participants prior to data collection. Participants were clearly informed about the purpose of the study, the voluntary nature of their participation, and their rights to decline participation, withdraw from the study at any time, or skip any question without any consequences to their care. All data were handled confidentially. No directly identifiable personal information was collected or retained.

## Results

### Sociodemographic characteristics and diagnostic profile

The study analyzed 165 vascular patients with a mean age of 49.6 years, comprising 53.3% males and 46.7% females, with the majority from the Addis Ababa, Oromia, and Amhara regions. Peripheral artery disease (PAD) was the most common diagnosis (47.3%), followed by carotid disease (20%) and various aneurysms (15.1%). The majority (69.7) were paying out of pocket money. The average hospital stay was 13.7 days. Among the 165 patients evaluated, 9.7% were former smokers, 1.8% were active smokers, and 88.5% had never smoked. Sixty-nine patients (41.8%) had at least one comorbidity, whereas 58.2% had none. Preoperative ASA classification revealed that most patients were ASA I–III (Table 1, Figure 1).

**Figure 1:**
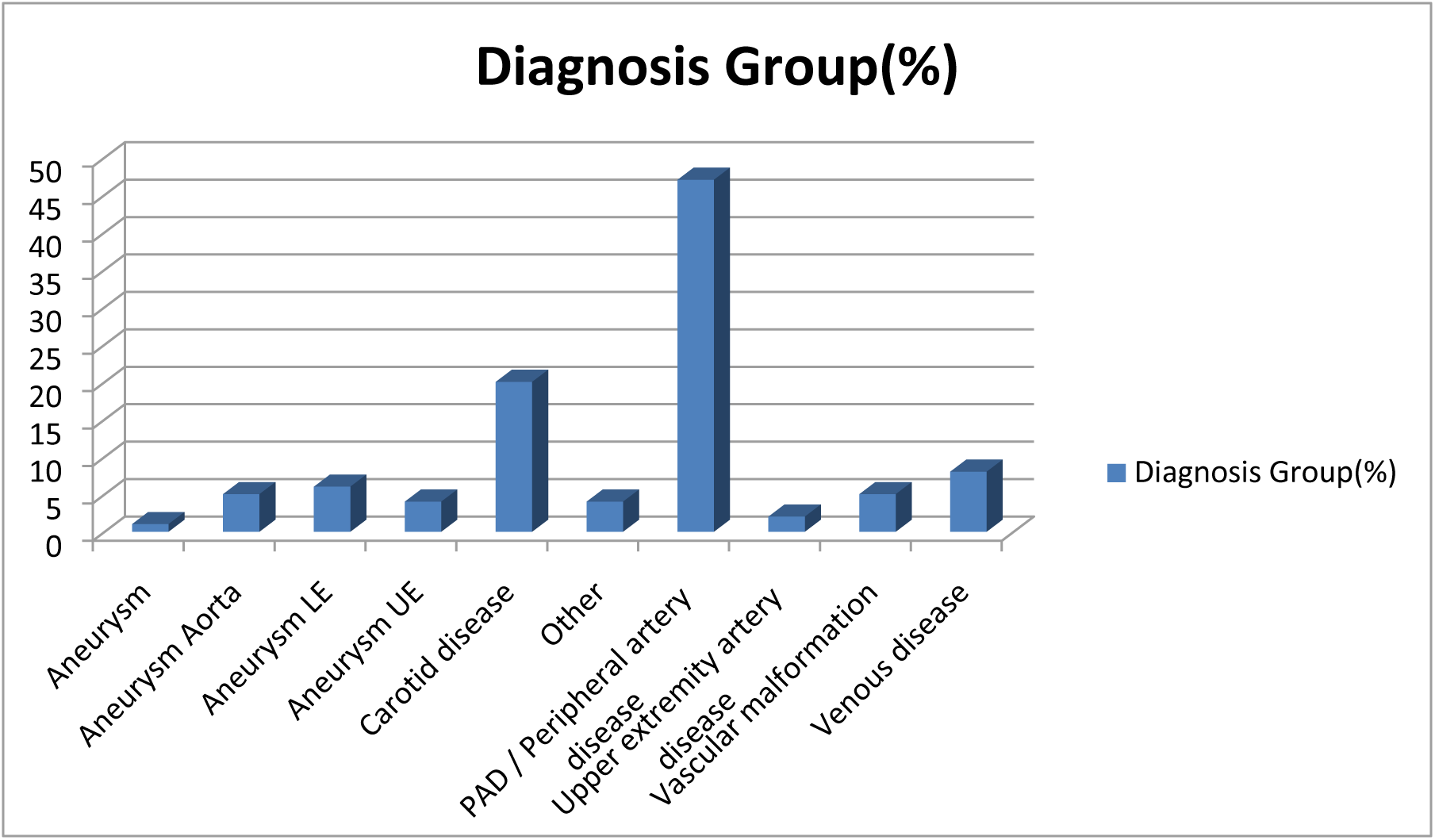
Diagnoses of patients

**Table 1:** Characteristics of the patients.

| Variables |  | Findings |
| --- | --- | --- |
| Age |  | Mean 49.6 (Min 17 & Max 87) |
| Gender | Male | 88 (53.3%) |
|  | Female | 77 (46.7%) |
| Smoking | Never smoked | 146 (88.5%) |
|  | Stopped smoking | 16 (9.7%) |
|  | Active smoking | 3 (1.8%) |
| Alcohol use | No | 152 (92.1%) |
|  | Yes | 13 (7.9%) |
| Comorbidities | No | 96 (58.2.0%) |
|  | Yes | 69 (41.80%) |
| Payment mode | Health insurance | 50 (30.3%) |
|  | Out of pocket | 115 (69.7%) |
| ASA classification | ASA I | 63 (38.2%) |
|  | ASA II | 45 (27.3%) |
|  | ASA III | 53 (32.1%) |
|  | ASA V | 4 (2.4%) |
| Mean hospital stay |  | Mean: 14 days (Min:2 days Max:50days) |

Compared with patients with other diagnoses, patients with peripheral artery disease and upper extremity artery disease had the highest average cost of investigation (Table 2).

**Table 2.** Average cost of investigations across diagnosis groups.

| Diagnosis Group | Average of Total Cost of Investigations (In Ethiopian Birr) |
| --- | --- |
| Aneurysm Aorta | 10859 |
| Aneurysm L.E | 10117 |
| Aneurysm U.E | 8223 |
| Carotid disease | 8885 |
| Other | 9048 |
| PAD(Peripheral artery disease) | 11866 |
| Upper extremity artery disease | 11738 |
| Vascular malformation | 8566 |
| Venous disease | 4191 |

Revascularization of the lower extremities was the most frequently performed procedure (35.8%), and 69.7% of the total procedures were high risk.

### Profile of all investigations

In general, preoperative investigations in vascular patients include imaging, laboratory tests and cardiac evaluation tests. No pulmonary function tests were performed in this study. CT was the most common imaging test (136/165, 82.4%), followed by ultrasound (74.5%). CBC was the most common laboratory test (100%), followed by renal function tests (RFT), electrolytes 97% and liver function tests (LFTs). (Table 3)

**Table 3:** Profile of investigations.

| Investigation | Done | Frequency | % done at private facilities(Outside the hospital) |
| --- | --- | --- | --- |
| CT scan | Yes | 136(82.4%) | 78.2% |
|  | No | 29(17.6%) |  |
| MRI | Yes | 4(2.4%) | 100% |
|  | No | 161(97.6%) |  |
| ULTRASOUND | Yes | 123(74.5%) | 30.1% |
|  | No | 42(25.5%) |  |
| ECG | Yes | 120(72.7%) | 38.3% |
|  | No | 45(27.3%) |  |
| ECHOCARDIOGRAM | Yes | 101(61.2%) | 47.5% |
|  | No | 64(38.8) |  |
| CXR | Yes | 33(20.0%) | 0.0% |
|  | No | 132(80.0%) |  |
| PFTs | Yes | 0(0%) | 0.0% |
|  | No | 165(100%) |  |
| CBC | Yes | 165(100%) | 0.0% |
|  | No | 0 (0%) |  |
| Blood Group | Yes | 158(95.8%) | 0.0% |
|  | No | 7(4.2%) |  |
| RFT | Yes | 163(98.8%) | 4.8% |
|  | No | 2(1.2%) |  |
| Electrolytes | Yes | 160 (97%) | 2.4% |
|  | No | 5(3.0%) |  |
| Calcium | Yes | 10(6.1%) | 30.0% |
|  | No | 155(93.9%) |  |
| Albumin | Yes | 18(10.8%) | 16.7% |
|  | No | 147(89.2%) |  |
| Coagulation | Yes | 147(89.1%) | 78.9% |
|  | No | 18(10.9%) |  |
| HIV Tested | Yes | 20(12.1%) |  |
|  | No | 145(87.9%) | 0.0% |
| **LFTs** | Yes | 159(96.4%) |  |
|  | No | 6(3.6%) | 1.9% |

### Imaging and cardiac test investigations

Regarding investigations, for 82.4% of patients, computed tomography (CT) scans were performed, with an average cost of 8,355 Ethiopian Birr (ETB). Among all the CT scans performed, 78.2% were performed in private diagnostic centers. Magnetic resonance imaging (MRI) was performed in 2.4% of the patients (mean cost 9,750 ETB) exclusively in private centers, and 74.5% of patients had ultrasounds (mean cost 523 ETB), with 30.1% performed in private centers. Electrocardiography (ECG), with an average cost of 513 ETB, was performed for 72.7% of the cases; 70.3% of the ECGs requested by surgeons and 20% of the ECGs were performed without indications.

Echocardiography, with an average cost of 732 ETB, was performed for 61.2% of the patients, with a nearly equal distribution between TASH (32.1%) and private health facilities (29.1%). A total of 43.5% of the echocardiograms were performed routinely without indications. Chest X-rays were performed for 20% of the cases and were performed in the hospital. The cost of chest X-ray was consistently 80 ETB (Table 4).

**Table 4.** Imaging investigations of patients, average costs and proportions performed at private centers.

| Investigation | % of patients with Investigations done | % of Investigation done in private | Average Cost in ETB | % of patients: Investigation was done not indicated |
| --- | --- | --- | --- | --- |
| CT Scan | 82.4% | 78.2% | 8,500 | 0.0% |
| MRI | 2.4% | 100% | 9,750 | 0.0% |
| Ultrasound | 74.5% | 30.1% | 523 | 0.0% |
| Chest X-ray | 20% | 0% | 80 | 0.0% |
| ECG | 72.7% | 38.3% | 513 | 20% |
| Echocardiography | 61.2% | 47.5% | 732 | 43.5% |

### Pulmonary function test (PFT)

Pulmonary function tests were not performed for any of the patients in our sample. Despite being indicated in 6 cases (3.6%). This finding suggests a gap in pulmonary system evaluation in vascular patients.

### Laboratory Investigations

CBC tests were provided for all patients and were routinely provided without indication in only 15 (9.1%) patients. The majority of CBC requests were made by surgeons (78.8%). All CBCs were provided by TASH, with a consistent cost of 50 ETB per test across all patients. A blood group determination test was performed for 95.8% (158/165) of the patients. The test was indicated for 148 patients (89.7%). A small portion (4.2%) did not perform this test. Surgeons and anesthesiologists were responsible for requesting all tests, while TASH provided almost all of them. The mean cost was approximately 30 ETB.

The majority of patients, 98.8% (163 out of 165), were tested. RFT was not indicated for 14.5% of all patients. The tests were primarily provided by TASH (94.5%), with a small portion performed at private centers (4.8%). The cost for RFTs varies widely, ranging from 100 to 800 ETB, with a mean of approximately 130.57 ETB. Electrolyte testing was performed in 97% of the patients. Although the test was indicated in approximately 68% of the cases, a notable 32% received it even when not indicated. Most requests were from anesthesiologists (61.8%), followed by surgeons (35.8%). TASH conducted nearly all the tests (97.6%). The mean cost was 447.19 ETB.

Only 6.1% of patients (10 out of 165) had calcium levels assessed, despite being indicated in 28% of patients. A significant proportion of calcium testing was performed at TASH (70%), and 30% was performed in private facilities. The average cost was 213.75 ETB, and most patients charged 185 ETB. The low rate of serum calcium testing explains this gap. Albumin testing was conducted in only 10.9% of patients, despite being clinically indicated in 53.9% of cases. Most albumin tests were provided by TASH (83.3%), and a small number were provided by private institutions. The average cost was 102 ETB, with a common charge of 50 ETB. A significant majority of patients (89.1%) did not have the test performed, which explains the gap in serum ALB testing.

A coagulation profile was completed for 89.1% of patients, with 86.1% having a clinical indication for it. The primary providers were anesthesiologists (61.8%) and surgeons (27.9%). The tests were performed mainly at private facilities (78.9%). The average cost was 508.76 ETB. Only 12.1% of patients underwent HIV testing, all of whom were treated at TASH and provided free of charge. LFTs were widely performed in 96.4% of patients, despite being clinically indicated in only 12.7% of patients. They were requested by surgeons, accounting for 88.5% of all LFT orders. Nearly all tests were performed at TASH, with costs averaging 150 ETB (Table 5).

**Table 5.**
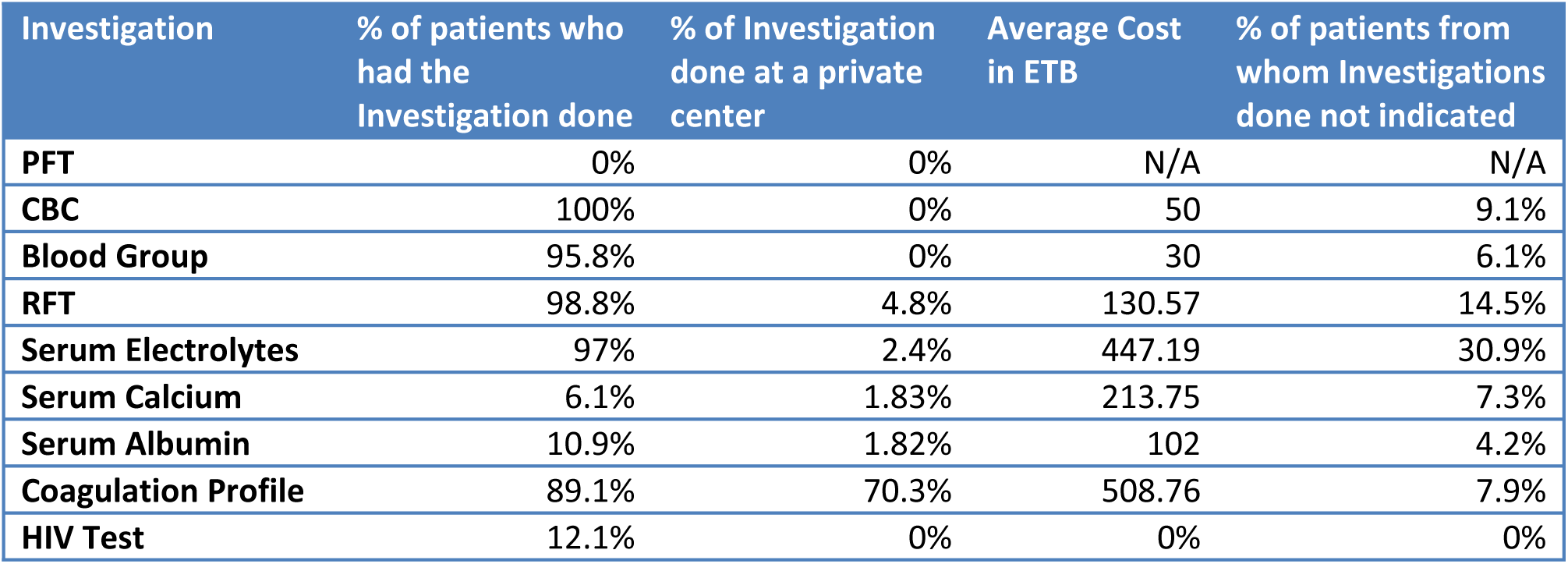
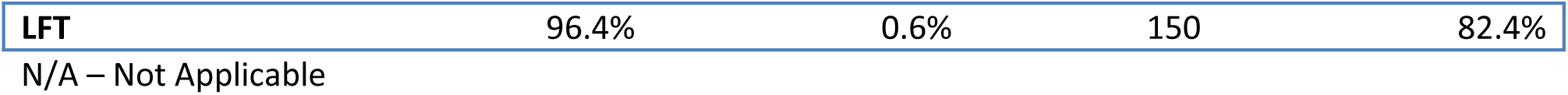
Laboratory investigations of patients, average costs and proportions performed at private centers.

| Investigation | % of patients who had the Investigation done | % of Investigation done at a private center | Average Cost in ETB | % of patients from whom Investigations done not indicated |
| --- | --- | --- | --- | --- |
| PFT | 0% | 0% | N/A | N/A |
| CBC | 100% | 0% | 50 | 9.1% |
| Blood Group | 95.8% | 0% | 30 | 6.1% |
| RFT | 98.8% | 4.8% | 130.57 | 14.5% |
| Serum Electrolytes | 97% | 2.4% | 447.19 | 30.9% |
| Serum Calcium | 6.1% | 1.83% | 213.75 | 7.3% |
| Serum Albumin | 10.9% | 1.82% | 102 | 4.2% |
| Coagulation Profile | 89.1% | 70.3% | 508.76 | 7.9% |
| HIV Test | 12.1% | 0% | 0% | 0% |
| **LFT** | 96.4% | 0.6% | 150 | 82.4% |

N/A – Not Applicable

Approximately 47.3% of patients received additional laboratory investigations, such as inflammatory markers and hepatitis viral markers. The cost of these tests ranged from 30 to 285 ETB, with a mean of 124.59 ETB.

### Investigation Costs

The mean total cost of imaging investigations was approximately 8,797 ETB, reflecting the use of advanced or private imaging services. Laboratory investigations cost an average of 1,328 ETB, ranging from 745 to 2,780 ETB. The average cost of investigations per patient was 9,717.40 ETB, with a median of 10,130 ETB.

The expense of CT scanning at TASH was increased by the cost of contrast bought in private pharmacies, and the average cost of contrast for one CT scan was approximately 5150 ETB. The average costs of investigations at private health facilities compared with TASH are shown in Table 6.

**Table 6:** Cost across private facilities vs TASH.

| Investigations | Average Cost at private in ETB | Average Cost at TASH in ETB |
| --- | --- | --- |
| **CT Scan** | 9,070 | 5,700 with contrast<br>550 without contrast |
| **MRI** | 9,750 | N/A |
| **Ultrasound** | 1,425 | 125 |
| **ECG** | 1,003 | 191 |
| **Echocardiography** | 972 | 186 |
| **Chest X-ray** | N/A | 80 |
| **PFT** | N/A | N/A |
| **CBC** | N/A | 50 |
| **ABO-Rh** | 30 | N/A |
| **RFT** | 563 | 100 |
| **Serum Electrolytes** | N/A | 450 |
| **Serum Calcium** | 185 | 300 |
| **Serum Albumin** | 50 | 300 |
| **Coagulation Profile** | 150 | 595 |
| **HIV Test** | 0 | N/A |
| **LFT** | 150 | N/A |

N/A – Not Applicable

### Affordability and Socioeconomic Factors—Financial Burden of Investigations

The mean monthly income was approximately 9,748 ETB, and the median income was 8000 ETB. However, it ranged widely up to 50,000 ETB. Notably, a significant number (32.7%) did not report income, underscoring socioeconomic data limitations.

### Gaps in Investigations

Our study analyzed disparities between what investigations were clinically indicated and what was actually provided for vascular patients in this study. Substantial gaps between the indicated and requested investigations among vascular patients were observed for serum calcium, pulmonary function tests (100% not performed) and serum albumin (89.5% not performed). HIV screening was also missed in 87.9% of the patients despite being free of charge.

### Cost of Unnecessary Investigations

Investigations that are ordered routinely without clinical indications represent a significant financial burden. More considerable expense was found with echocardiography, with a mean cost of 197.52 ETB, and 26.7% of patients underwent this test without clinical indication. Total Unnecessary Investigation Cost: This cost was an average of 648.99 ETB per patient, with the highest cost reaching 3,565 ETB (Table 7).

**Table 7.** Cost of unnecessary investigations.

| Test Type | Total Test Done | Not Indicated | % Not Indicated | Total Cost (ETB) |
| --- | --- | --- | --- | --- |
| ECG | 120 | 24 | 20.00% | 20051.4 |
| Echocardiography | 101 | 44 | 43.56% | 32208 |
| CBC | 165 | 15 | 9.09% | 750.75 |
| ABO-Rh | 159 | 11 | 6.92% | 300.30 |
| RFT | 164 | 24 | 14.63% | 3,649.80 |
| Electrolytes | 161 | 50 | 31.06% | 22,948.85 |
| Calcium | 10 | 0 | 0.00% | 0 |
| Albumin | 18 | 8 | 44.44% | 849.75 |
| Coagulation Profile | 148 | 13 | 8.78% | 5,430.15 |
| LFT | 160 | 138 | 86.25% | 20,101.92 |
| Total |  |  |  | 106,675 |

### Hospitalization Costs

The average cost of hospital stay (without insurance consideration) was 4,344 ETB, ranging up to 15,900 ETB. When accounting for insurance or cost coverage, the average real cost to patients dropped to 2,906 ETB. Notably, 29.7% of patients had zero out-of-pocket expenses for hospitalization.

The total cost of combining investigations and hospital stay (with insurance considered) averaged 12,623.73 ETB per patient, ranging from 125 to 35,400 ETB. This represents a significant expenditure for most patients.

### Associations between the Cost of Investigations and Patient Variables

Linear regression was conducted to explore the relationships between patient demographics, clinical factors, and both the number and cost of investigations. The dependent variable was the total cost of investigations. The linear regression analyses revealed several significant predictors of total cost of investigations. Age was positively associated with increased total cost, with each additional year linked to a 78.2-unit increase in cost (p < .001). A higher classification of ASA classes also significantly predicted increased costs (B = 1473.97, p < .001), suggesting that patients with poorer preoperative health status incurred greater expenses.

Comorbidity was another strong predictor, with a B value of 2108.92 (p < .001), indicating that patients with more comorbid conditions had significantly higher costs. Similarly, longer hospital stays were associated with increased costs (B = 107.06, p < .001). On the other hand, the diagnosis group and mode of payment were both significantly associated with lower total costs (p = .037 and p = .032, respectively). The type of procedure and use of alcohol were not statistically significant predictors of cost, with p values above 0.05 (Table 8).

**Table 8.**
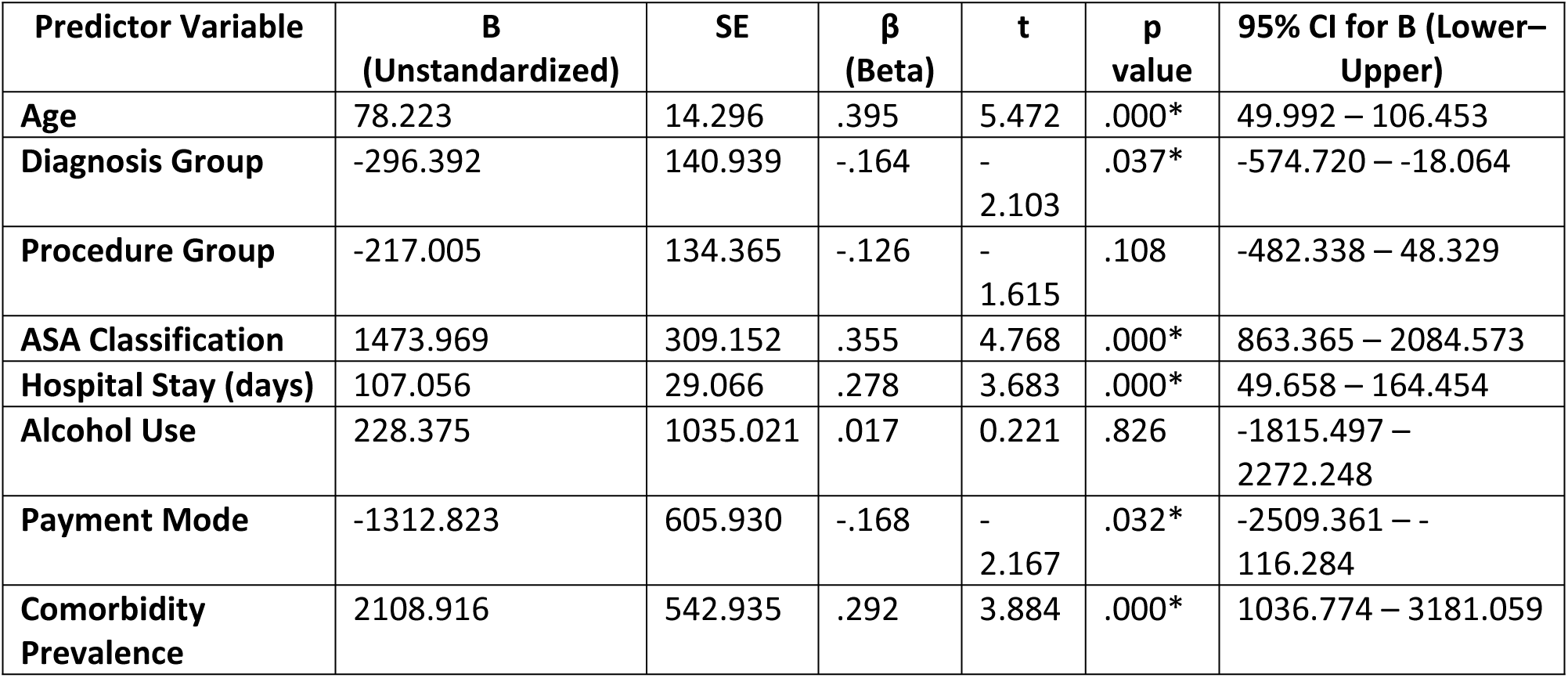
Linear regression analysis between total investigation cost and patient variables.

| Predictor Variable | B<br>(Unstandardized) | SE | $\beta$<br>(Beta) | t | p<br>value | 95% CI for B (Lower–Upper) |
| --- | --- | --- | --- | --- | --- | --- |
| Age | 78.223 | 14.296 | .395 | 5.472 | .000* | 49.992 – 106.453 |
| Diagnosis Group | -296.392 | 140.939 | -.164 | -2.103 | .037* | -574.720 – -18.064 |
| Procedure Group | -217.005 | 134.365 | -.126 | -1.615 | .108 | -482.338 – 48.329 |
| ASA Classification | 1473.969 | 309.152 | .355 | 4.768 | .000* | 863.365 – 2084.573 |
| Hospital Stay (days) | 107.056 | 29.066 | .278 | 3.683 | .000* | 49.658 – 164.454 |
| Alcohol Use | 228.375 | 1035.021 | .017 | 0.221 | .826 | -1815.497 – 2272.248 |
| Payment Mode | -1312.823 | 605.930 | -.168 | -2.167 | .032* | -2509.361 – -116.284 |
| Comorbidity Prevalence | 2108.916 | 542.935 | .292 | 3.884 | .000* | 1036.774 – 3181.059 |

## 5. Discussion

This study highlighted the investigations ordered for preoperative surgical patients and the associated costs at TASH. The burden of unnecessary preoperative investigations is amplified by limited public resources and out-of-pocket payments (30). In this study, the out-of-pocket expenditure was 69.7%. This is greater than the estimation reported in studies performed in Ethiopia, which revealed that one-third of total health expenditures come from out-of-pocket payments, with inadequate financial protection for surgical patients (28,29). Mollalign et al. reported that 13.9% of the preoperative cost burden in Gondar was due to unnecessary tests (24), whereas a recent study at TASH reported that more than 30% of tests ordered preoperatively lacked clinical indications (25).

In this study, imaging tests increased the total cost. CT scans and echocardiography followed by excessive routine laboratory tests were performed. Notably, while some high-value investigations, such as pulmonary function tests and serum ALB, have been underutilized despite clinical indications, others have been performed excessively. PFTs, which were never performed, highlight systemic gaps during the preoperative evaluation. These patterns are concerning given that only a small proportion of the results influence patient management, which is consistent with findings by Lakomkin et al. and Admass et al. (12,16). Pollard et al. previously demonstrated that outpatient preoperative evaluations could reduce hospital length of stay and cost without compromising care quality (9).

The analysis of preoperative evaluation in vascular surgery patients at TASH revealed excessive utilization of investigations and unneeded costs driven by routine testing. Nearly all patients underwent basic laboratory examinations (e.g., 100% had CBC, 98.8% RFT, 97% electrolytes), and a majority received imaging (CT 82.4%, ultrasound 74.5%, ECG 72.7%). However, many tests lack clear indications; for example, only 64.2% of ECGs and 36.4% of echocardiograms are clinically justified. These findings mirror global reports of wasteful preoperative testing. A study in the Caribbean reported that 64% of tests were unnecessary, resulting in major financial losses (11). Lakomkin et al. reported that routine preoperative laboratory tests in orthopedic trauma patients did not affect treatment outcomes (12). Similarly, Vogt et al. and Obasuyi and Antwi-Kusi reported that more than 70% of tests ordered by surgeons were deemed unnecessary, contributing to significant cost burdens (13).

In this study, CT scans were used as a single investigation to increase the total cost and were largely performed at private centers. This aligns with the literature noting the high costs of advanced imaging (17, 18). Rubin et al. reported that 10–30% of imaging tests were performed without clinical justification, leading to unnecessary radiation and financial costs (20). Shaw et al. demonstrated that selective preoperative stress testing can significantly reduce costs in vascular procedures (21). In the United States, routine preoperative testing is estimated to cost over $30 billion annually (22). Harris et al. reported similar findings within the Virginia health system, where nearly half of the preoperative test costs were from low-value care (23).

However, in our study, imaging investigations were performed with clinical indications. Even though they were indicated, they still raised financial burdens to the patients, as more than half of our study participants mentioned that they could not afford the investigations. This could be attributed to the high cost of healthcare, the unavailability of functional imaging investigation tools at public hospitals and the low socioeconomic status of the population.

With respect to laboratory investigations, while routine tests such as CBC and blood group typing are almost universally indicated and inexpensive, other tests have shown overuse. Electrolyte panels were performed in 97% of patients, despite being clinically indicated in only 68% of cases, whereas tests such as LFTs were widely overused. These findings are consistent with those of Admass et al., who reported that 60–70% of routine laboratory tests were unnecessary and contributed to elevated healthcare costs (16). Akula and Vijaynagar similarly reported the potential for substantial savings by limiting routine blood groups and cross match orders (15). Although not all patients in our study were tested for HIV, studies have shown the impact of early HIV testing to prevent complications in elective surgery patients (31). This is possibly due to the lack of local guidelines on HIV screening in elective surgical patients in TASH.

This study also quantified the financial burden of unnecessary preoperative investigations, revealing a substantial avoidable cost to patients. On average, patients incurred 648.99 ETB in unnecessary test-related expenses, with some paying as much as 3,565 ETB. The largest contributors were echocardiography and electrolyte panels, followed by LFTs and ECGs. These findings parallel international data that estimate that up to 60–70% of routine preoperative tests are not clinically necessary and lead to excessive costs without improvement in patient outcomes (13,16,19,22). Prior studies have also reported high rates of test overuse: a study in Gondar attributed 13.9% of investigation costs to unnecessary testing (24), and another study at TASH reported that more than 30% of elective surgery patients underwent unnecessary preoperative tests (25). Given the out-of-pocket expenditure burden (28, 30) and the low income of patients in general, these avoidable costs likely exacerbate the cost of surgical care and may lead patients to catastrophic health expenditures.

The analysis of patient variables in our study revealed significant factors associated with increased investigation costs: age, diagnosis, hospital stay, ASA class, payment mode and comorbidities. A linear relationship was observed between patient age and total investigation cost (p < 0.001), with each additional year of age contributing to an estimated 78.2 ETB increase. This aligns with prior findings that older vascular patients, who often have multiple risk factors, require more extensive preoperative evaluation (3,4). Additionally, patients with comorbidities incurred an average additional cost of approximately 2,109 ETB (p < 0.001), supporting the literature that highlights comorbid conditions as drivers of increased testing and healthcare utilization (6,7). In contrast, no significant associations were observed between investigation costs and procedure type or alcohol intake. These results suggest that clinical complexity rather than procedural complexity is the principal determinant of preoperative investigation costs, which is consistent with existing data emphasizing the need for risk-based testing (21,26).

In this study, the mean monthly income was approximately 9,748 ETB, whereas the average cost of preoperative investigations per patient was 9,717.40 ETB, which could highlight the potential impoverishment that a vascular patient would face if he was likely to undergo surgery.

## Conclusion and Recommendations

### Conclusion

This study demonstrated that preoperative investigations in vascular surgery at TASH are characterized by high rates of overinvestigation. Certain laboratory tests are frequently ordered without clinical indications, resulting in significant direct costs to patients, and many of them already face substantial financial vulnerability. Preoperative tests should be ordered on the basis of patient history, comorbidities and physical findings to avoid financial risk that can result from unneeded tests requested routinely. By implementing this approach, it is possible to preserve healthcare resources and protect vascular patients from catastrophic health expenditures in TASH. These findings are not only relevant for TASH but also offer important insights for broader surgical systems across low- and middle-income countries facing similar challenges.

### Recommendations

The implementation of a locally agreed-upon protocol for presurgical investigations of vascular patients considering each patient condition could help limit and decrease the number of costly unneeded tests that are routinely provided. The avoidance of imaging and laboratory tests, such as coagulation profiling, limits the number of expensive tests that are performed outside the hospital and are not affordable for most patients. A prospective study is needed to investigate the factors contributing to requests for routine preoperative investigations for vascular patients. A cost-effective preoperative investigation for vascular patients is also needed.

The ministry of health can work to increase the number of community health insurance **members,** especially for **the** old population and people with chronic diseases, as they face a financial burden for preoperative investigations when they need surgery and for costs that come from care at private facilities. Financial protection strategies for health care should be reinforced to promote timely access to safe and affordable surgery, as many patients may not afford the cost of surgery considering the high cost of preoperative investigations through making necessary investigations available at hospitals where costs could be covered by health insurance.

### Limitations of the Study

This study provides primary data on investigation costs for vascular patients undergoing surgery and can serve as a reference for further studies. This was a single-center study, and the results may not be generalizable to the whole population. There was an arrangement order of papers in patients’ files submitted for archival, which made it difficult to find records such as operation notes and investigation results.

## Data Availability

The datasets used in this study are available from the corresponding author upon request.

## List of abbreviations

ASA: American Society of Anesthesiologists
aPTT: activated partial thromboplastin time
CT Scan: Computed tomography scan
ETB: Ethiopian Birr
ECG: Electrocardiography
ECHO: echocardiography
INR: International normalized ratio
MRI: Magnetic resonance imaging
PAC: Preanesthesia checkup
PT: Prothrombin time
SPSS: Statistical Package for the Social Sciences
TASH: Tikur Anbessa Specialized Hospital
USD: United States Dollar

## Availability of data and materials

The datasets used in this study are available from the corresponding author upon request.

## Authors’ contributions

KD wrote the proposal, ST collected the data, and AM analyzed the data. KD and AM wrote the final manuscript. TD reviewed and provided directions during the write-up process.

## Declaration of Generative AI and AI-assisted technologies in the writing process

During the preparation of this work, the author used Curie to improve the language. After using this tool/service, the author reviewed and edited the content as needed and takes full responsibility for the content of the published article.

## Disclaimer

The views and opinions expressed in this article are those of the author/authors and are the product of professional research. They do not necessarily reflect the official policy or position of any affiliated institution, funder, agency, or publisher. The authors are responsible for this article’s results, findings, and content.

